# EMS responses to hospice patients: a qualitative study

**DOI:** 10.1101/2022.04.08.22272501

**Authors:** Whitney Kukol, Christopher W. Ryan

## Abstract

In the palliative care community, multiple emergency department visits and hospitalizations are considered an indicator of poor-quality end of life care. There is substantial research on the events surrounding ED visits, and subsequent hospitalizatons, by hospice patients. It is important however to understand that these hospital arrivals often begin with an ambulance response to the home. Emergency medical services (EMS) responses to hospice patients have been less well-studied. We conducted a retrospective study of 170 consecutive electronic patient care reports produced contemporaneously by EMS providers in several US counties during the normal course of their responses to hospice patients. We summarized descriptive epidemiology of the patients and of the incidents, and we explored the provider narratives for recurring themes. To our knowledge this is the first study to explore the contemporaneously-documented on-scene circumstances of pre-hospital care for patients enrolled in a hospice program.

The median patient age was 76. Cancer, chronic lung disease, and heart failure were the most common hospice-qualifying diagnoses. Transportation of the patient from the scene occurred in 111 cases (65.3%). Consistent with previous studies based on interviews of EMS providers, the most common chief complaints included pain, dyspnea, and altered mental status. EMS was frequently called for confirmation of death, a finding not previously reported.

Transitions of care, such as from curative medical care to hospice care, and from hospital to home, were a recurring theme. Gaps in care around these transitions were often evident, with EMS being called upon to fill them. Care transitions around weekends were prominent. Also emerging from the narratives was the concept that hospice enrollment is not a single event but rather a step-wise process; we developed a preliminary coding taxonomy to classify those enrollment stages.

The hospice-EMS interface can be complex. Although all care by EMS providers is with the literal or implied consent of the patient, the arrival of an ambulance to a scene where emotions are running high carries a certain momentum and could lead to a cascade of interventions that, in retrospect and given other options, the patient may not have wanted. A better understanding of the hospice-EMS interface might illuminate changes that could be made to improve it, with a goal of ensuring that hospice patients and their families receive emergency medical services when they need and can benefit from them but receive other, non-EMS, services when those are more suitable to their needs. It might also improve the efficiency of an already overburdened EMS system.

## 2 Background

The purpose of hospice care is to help patients and their loved ones along the journey to death, when that death can no longer be avoided or postponed by available medical treatments. Hospice care provides compassionate end of life care in the comfort of a patient’s home or a home-like facility. Objectives include relief of physical, emotional, and spiritual distress in patients and their loved ones.

However, this journey is often punctuated by distressing events that patients/families are sometimes not expecting: new symptoms, sudden worsening of existing symtpoms, or sudden decline in function. Although members of the hospice team make regular visits to the patient’s residence and are on-call to respond 24 hours a day,^1^ patients enrolled in hospice still sometimes go to emergency departments (ED). In the palliative care community, multiple emergency department visits and hospitalizations are considered an indicator of poor-quality end of life care.^2^ There is substantial research on the events surrounding ED visits by hospice patients. Several retrospective chart review studies, in different countries, reported that the most common presenting ED complaints among hospice patients were pain, dyspnea, vomiting, altered mental status, fever/chills, and general weakness.^3–7^ These ED visits are often considered avoidable, and strategies to predict them and reduce their occurrence have been explored.^6–9^ For unavoidable ED visits by hospice patients, efforts to improve the patient and family experience have been described.^10^

The existing literature on ED utilization by hospice patients largely begins at the door of the ED. But patient care often starts “upstream,” in the prehospital setting, when emergency medical services (EMS) is called to the patient’s location. Thus it is important to understand how, when, and why patients enrolled in hospice care utlize EMS.

The hospice-EMS interface can be complex. Although all care by EMS providers is with the literal or implied consent of the patient, the arrival of an ambulance to a scene where emotions are running high carries a certain momentum and could lead to a cascade of interventions, including tranpsport to an ED, that, in retrospect, the patient may not have wanted. This “cascade effect” is well-recognized in medicine.^11^ On the other hand, some EMS responses to hospice patient might relieve distressing symptoms, enhance patient autonomy, and restore their homeostasis in their home—all goals of hospice care. For example, consider a non-transport call for “lift assist,” to return a fallen patient to their bed or chair safely and comfortably. As another example, EMS treatment and transport for a coincidental acute medical problem, unrelated to the hospice-defining diagnosis, might enable the patient to return home in comfort for the remainder of their life expectancy.

Understanding the phenomenon of EMS responses to patients enrolled in hospice programs could support patient- and family-centered end-of-life care, improve the working interface between EMS systems and hospice programs, and optimize utilization of EMS resources. Numerous studies using individual interviews, focus groups, and surveys have explored the recollections and perceptions of hospice staff,^12^ family members,^13^ and EMS providers^14–23^ of the role of EMS and hospitalization in palliative care. (It should be noted that terms like “palliative care” are sometimes used loosely in the literature, and it may not be limited to patients formally enrolled in a hospice program.) Some common themes emerged from these interviews.

The EMS providers collectively described: (1) EMS responses to patients in palliative or hospice care are commonplace, (2) an ambulance repsonse to a patient in palliative or hospice care is often the initial catalyst for a sequence of events that the patient may not want—the “cascade effect” mentioned above, (3) EMS education programs typically provide little or no training specific to this situation, and (4) the structure of the EMS system and its protocols can impede in-home palliative care.

Hospice personnel collectively related: (1) not all patients and families enrolled in hospice programs truly embrace this care model, (2) they can become frightened and call for an ambulance when death is imminent or when they are surprised by distressing symptoms, and (3) response times by EMS units can be shorter than those of hospice personnel.

In addition to voicing recollections very similar to those of hospice personnel, primary caregivers (usually family members) also described that they value continuity with the care team (often at the hospital) that they have become familiar with thoughout their loved one’s illness.

While these interview-based studies have yielded many insights, it would be worthwhile to try to understand the real-time details of events at the scene of EMS responses to hospice patients. To fill that research gap, we conducted a retrospective study of the electronic patient care reports produced contemporaneously by EMS providers during the normal course of their responses to hospice patients. We summarized descriptive epidemiology of the patients and of the incidents, and we explored the provider narratives for recurring themes.

## 3 Methods

The setting for this study is a multi-county EMS Region (the “Region”) upstate New York, with a combined population of about 300,000. The Region comprises rural and suburban areas, a number of villages, and two small cities. It is served by approximately 77 EMS agencies, of various structures: commercial, volunteer, fire-service-based, transporting, non-transporting, advanced life support (ALS), and basic life support (BLS). The system offers a tiered response, in which ALS is universally available, either for initial response or to join a BLS unit at the scene or in transit when needed.

The Region is served by several hospice programs. Their geographic areas of operation generally do not overlap. It would be rare for a patient in the Region to have a choice between hospice programs; patients would generally enroll in the program serving the location of their residence. In this study, we made no distinction between the the hospice programs, considering them collectively to be “hospice care.” EMS agencies with a total call volume during the study period of less than 50 were excluded, leaving 63 agencies that were invited to participate by allowing their data to be used. Eleven agencies agreed. These eleven comprise the largest and busiest agencies, and together they account for over 85% of total Regional call volume.

The Regional Emergency Medical Services Council (REMSCO) maintains an electronic database of patient care reports (ePCR) from all EMS agencies in the Region. The database complies with the National Emergency Medical Services Information System (NEMSIS) standards (http://www.nemsis.org/.) Information about incidents and patients is entered into the database by EMS crews in real time or nearly so. The Regional Emergency Medical Advisory Committee (REMAC)—a physician subcommittee of the REMSCO–is charged with stewardship of the database and approved its use for this study. The SUNY Upstate Medical University Institutional Review Board also approved the study.

EMS reports from participating agencies, generated between 1 January 2013 to 30 June 2016 inclusive, in which the word “hospice” appeared in the narrative, were retrieved. The collected data included: patient age, day of the week of EMS response, time of day of EMS response, and the EMS provider’s narrative (usually a short paragraph of text.) As required by the Institutional Review Board and federal privacy regulations, all patients over age 89 years were collapsed into a single age category. The ePCR database records age in two fields: an “age unit” field and a numeric field indicating the patient’s age in those units. All records with “age unit” other than “years” (e.g. days, weeks, or months) were excluded. This has the effect of excluding data from infants; an infant enrolled in hospice care is considered to be an unlikely scenario.

To protect privacy, where frequencies of events, conditions, or circumstances are lower than 5, they are presented as “fewer than 5.” The detailed narrative documentation in a pre-hospital patient care report could be particularly sensitive information. Especially in small communities, innoccuous single pieces of information might, in combination, inadvertantly disclose a patient’s or healthcare worker’s identity. Therefore, in the illustrative text segments presented here, potentially identifying information is replaced with empty square brackets. Textual material ommitted for brevity is replaced with ellipses. Comments from the authors to clarify jargon are enclosed in curly braces. Spelling and grammar errors in illustrative text segments have not been corrected.

## 4 Results

### 4.1 Characteristics of patients

The query of the ePCR database yielded 198 records. Twenty-eight were excluded because upon qualitative review of the narratives there was documentation that they were interfacility transports or calls to return patients home from a hospital. Thus the analytical dataset consisted of 170 records, each documenting the care of one patient by one EMS agency during one incident response.

Our primary purpose was to understand the events and dynamics surrounding EMS responses to hospice patients, based mainly on a qualitative review of EMS provider narratives for recurring themes. Nevertheless, some descriptive epidemiology of the incidents provides some useful context.

The median patient age was 76, with half of patients being between 64 and 85. For privacy purposes, ages over 89 years were counted as a group; there were 24 such patients.

When the reason or diagnosis for which a patient was enrolled in hospice was mentioned in the narrative, this was coded into broad categories that emerged from the coding process; these are shown in Table 1. In some cases, no reason for hospice enrollment was documented. This was sometimes found in cases in which hospice enrollment was under consideration or underway (see Section 4.3.1.

**Table 1:**
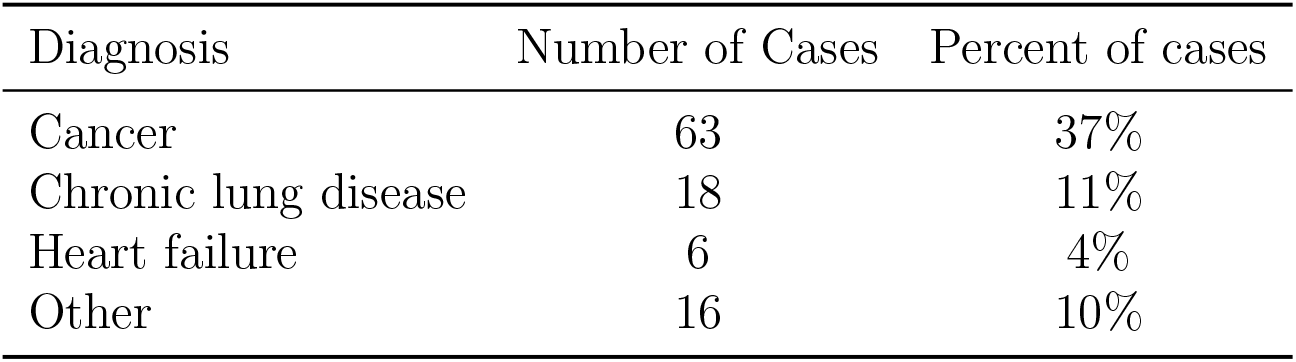
Hospice-defining diagnoses mentioned in EMS provider narratives in responses to hospice patients. Diagnoses mentioned in fewer than 5 cases are combined into “Other.” Cases in which the reason for being enrolled in hospice was not documented are not included in the table.

Patient chief complaint represents the reason(s) they or a bystander called EMS, or their main concern(s), as expressed to the EMS crew upon their arrival. Chief complaints are rarely the same as the underlying or “background” hospice-defining diagnoses. The nature of the chief complaints appearing in the narratives was assessed by qualitative review of the narratives and grouped into broad categories that emergend from our analysis; the frequencies of the categories are shown in Table 2. The chief complaint categories are not mutually exclusive; it is possible that more than one chief complaint is documented in a single case.

**Table 2:** Chief complaints (i.e. reasons for calling EMS as expressed to the arriving EMS providers) mentioned in narratives in EMS responses to patients enrolled in hospice. All chief complaints occuring in fewer than 5 cases are combined into “Other.”

| Chief complaint | Number of cases | Percent of cases |
| --- | --- | --- |
| Pain | 38 | 22% |
| Fall | 31 | 18% |
| Difficulty breathing | 28 | 17% |
| Altered mental status | 19 | 11% |
| Confirmation of death | 12 | 7% |
| Difficulty getting around <sup>1</sup> | 10 | 6% |
| Poor oral intake | 6 | 4% |
| Generalized weakness | 5 | 3% |
| Nausea/vomiting | 5 | 3% |
| Constipation | 5 | 3% |
| Other | 9 | 8% |

<sup>1</sup> “Difficulty getting around” means that the patient was unable to get from where they were to where they wanted to be. This is sometimes referred to as needing “lift assist.” Sometimes it is subsequent to a fall.

### 4.2 Characteristics of incidents

#### 4.2.1 Timing of incidents

The distribution of the days of the week and the times of the day of the incidents are shown in Figures 1 and 2. The modal day in the sample was Friday, and the peak time was centered around 1000 hr, with a heavy tail into the afternoon.

**Figure 1:**
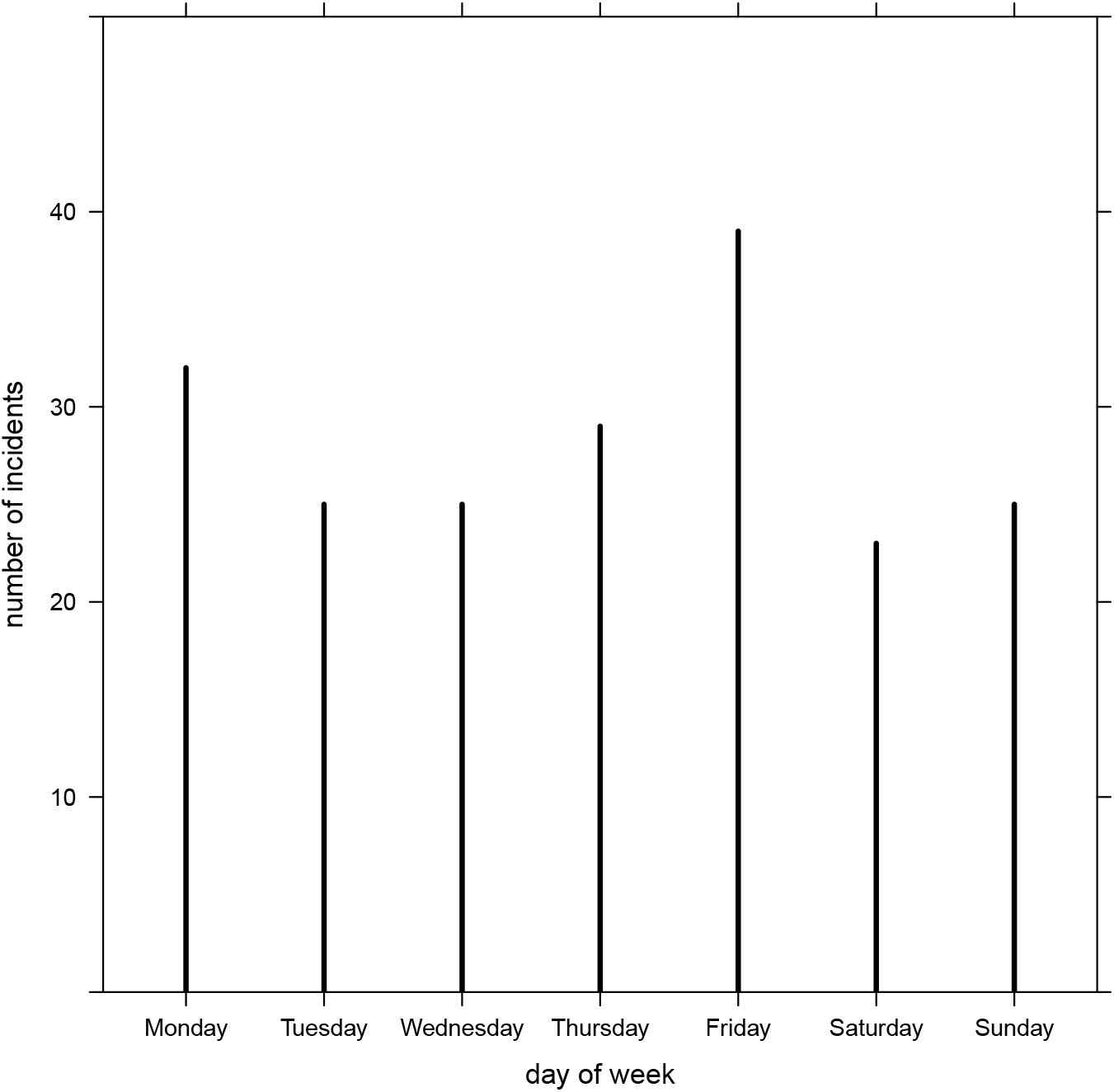
Days of the week on which EMS responses to hospice patients occurred. Weekdays are shown in order beginning at the left, which the weekend (Saturday and Sunday) is shown at the far right. No weekday was particularly prominent.

**Figure 2:**
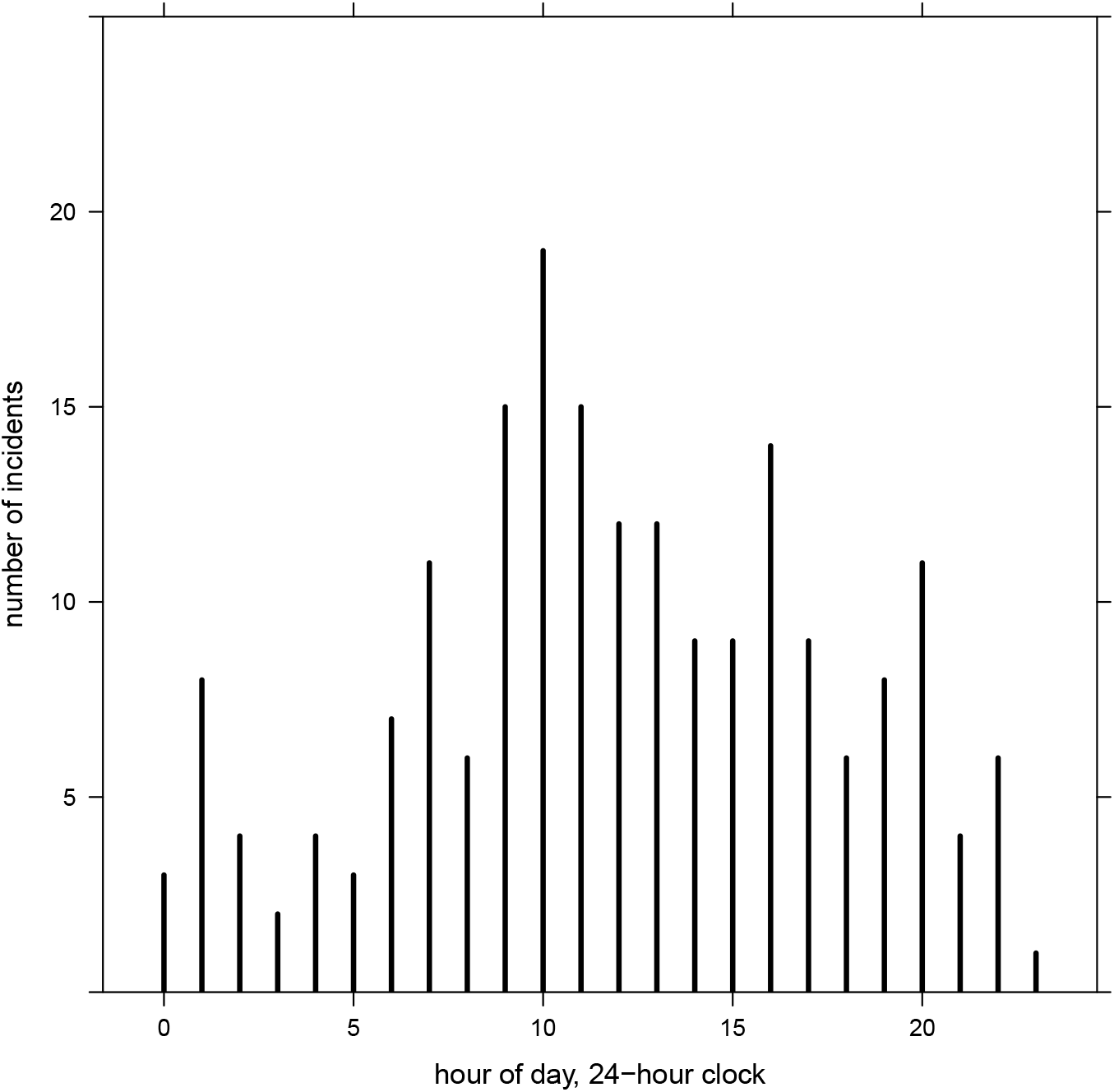
Time of day at which EMS responses to hospice patients occurred. Times are shown by the hour on a 24-hour clock.

#### 4.2.2 How EMS came to be called

The process that leads to a decision to call EMS can be complex. In our sample, EMS was called to the scene by a variety of people or agencies. Most frequently, of course, the patient or a family member called. In 17 cases (10%), there was documentation that hospice personnel called or advised the patient/family to call. Rarely, another agency made or recommended the call to EMS; these included physicians, facility (e.g. nursing home) staff, and in one case funeral home personnel (who were not at the scene). Hospice personnel were also called, or were already on scene, in 21 (12%) and 23 cases (14%) respectively. In 7 cases (4%) there was documentation that callers could not reach the hospice program or were disatisifed with the timeliness of their response, leading them to then call EMS.

#### 4.2.3 Events at the scene

Other people or agencies were often present at the scene, as shown in Table 3. No narratives documented that funeral home personnel or clergy were present at the scene.

**Table 3:** Frequency of cases in which various people and agencies were present at the scene of an EMS response to a hospice patient. “Home health aide” means a trained person employed to help care for the patient but not specifically documented as being affiliated with a hospice program. “Other” is usually in the context of a response to an institution with its own staff, such as a nursing home.”

| Person or agency | Number of cases | Percent of cases |
| --- | --- | --- |
| *Family members and friends* |  |  |
| unspecified relative | 40 | 23.5% |
| spouse | 18 | 16.5% |
| child | 21 | 12.4% |
| sibling | fewer than 5 |  |
| friend | fewer than 5 |  |
| parent | fewer than 5 |  |
| *Agency representatives* |  |  |
| hospice | 23 | 13.5% |
| law enforcement | 22 | 12.9% |
| home health aide | 10 | 5.9% |
| other | 6 | 4.7% |

A variety of actions by EMS personnel were documented in the narratives. Transportation of the patient from the scene occurred in 111 cases (65.3%). The destination was almost always one of the local hospital emergency departments, except in the rare instances of scheduled transport to a medical appointment (two cases). Other documented actions by EMS personnel are shown in Table 4. Other than oxygen, inhaled albuterol (with or without ipratropium) was the most frequently-administered medication documented in the narrative. In four cases, the narrative described administration by EMS of parenteral opioids for pain.

**Table 4:** Actions documented in EMS narratives during responses to hospice patients. “patient movement or lift assist” applies only to non-transported patients; in the case of transport from the scene, this is assumed.

| Action taken | Number of incidents | Percent of incidents |
| --- | --- | --- |
| EKG or cardiac monitor | 51 | 30% |
| vascular access | 45 | 25.5% |
| oxygen administration | 42 | 24.7% |
| medications other than oxygen | 19 | 11.2% |
| patient movement or lift assistance | 11 | 6.5% |
| ventilation | fewer than 5 | 2.4% |
| chest compressions | fewer than 5 | 1.8% |
| endotracheal intubation | fewer than 5 |  |

#### 4.2.4 Disposition

The narratives contained evidence of the patient being transported from the scene in 65% of cases, evidence of no transport in 11%, and evidence of refusal of medical assistance (RMA) in 8%. In 12% there was documentation that someone at the scene (e.g. patient, family, hospice personnel) specifically requested that the patient be transported. Conversely, in 4% a specific request *not* to tranpsort the patient was documented.

### 4.3 Emergent themes

#### 4.3.1 Transitions of care

Transitions of care and handoffs occur both within and between institutions, agencies, and care settings. The risks presented by care transitions have become an important topic in the healthcare safety literature,^24–26^ and this emerged as a recurring theme in our study as well.

It became clear that enrollment in hospice was not an event, or a binary variable, but rather a series of transitions from one state to another, evolving over time. The stages entailed increasing commitment to the idea of hospice care, reminiscent of the stages of change in the Transtheoretical Model,^27^ often outlined as precontemplation, contemplation, preparation, action, maintenance, and termination. We created the following coding taxonomy to classify the enrollment stages documented in the narratives. Frequencies and illustrative text segments are shown.

**considering enrollment** Enrollment in hospice has been discussed between the parties (usually patient, family, and physician) but no decision has yet been reached. Documentation of this stage was found in 7 cases (4%).

**intending to enroll** The patient has decided to enroll in hospice, but this has not yet been initiated. Documentation of this stage was found in 7 cases (4%).

**enrollment underway** The enrollment process has begun but is not yet complete. Documentation of this stage was found in 12 cases (7%).

**enrolled** The patient is enrolled in hospice care. Documentation of this stage was found in 104 cases (61%).

**recently enrolled** A sub-type of “enrolled,” arbitrarily defined as evidence of enrollment in the previous three days, or that the patient is enrolled but the initial caregiving visit (in contrast to a pre-enrollment assessment visit) has not yet occurred. There were 7 such cases (4%).

**formerly enrolled** Patient has been in hospice at some point but is not enrolled at the time of the EMS response. There were fewer than 5 such cases.

Transition from one enrollment stage to another, often accompanied by a transition from hospital to home, can risk gaps in care, which EMS may be called upon, by a distressed patient or family, to fill. EMS narratives from responses to several patients who were intending to enroll or whose enrollment was underway are illustrative:

> Dispatched to [] for possible unattended {“possible unattended” appears frequently in the narratives; it is professional jargon for an unattended death}. Upon arrival, was met by pt’s family at door with valid DNR. Pt’s family stated pt has a history of [] and was set to be put on hospice next week
>
> {was} … [] restless Breathing problems … told that home health nurse had been at residence administered P.O. [] an departed residence presently Hospice nurse in residence verbalize pt. is “not under their service as of present.” [family member] states [] has been attempting to have [patient] placed into hospice.
>
> Called for a reported Sick Person at []. On arrival, found a [] patient weighing 90 KG. Chief complaint of Increase in lethargy. Events surrounding incident: [family member] states that the Pt was evaluated at [] states that they are to meet to setup hospice care on Friday. However, the [family member] states that the Pt has been increasingly more lethargic [], and feels [] needs help with the Pt around the house.

One patient/family in the process of enrollment in hospice saw EMS as the route to evaluation by a physician that they were told was required for enrollment into hospice:

> Family stated they are in the process of having the patient placed into Hospice Care. Family stated they called 911 today because they were advised the patient needed to be evaluated by a physician before being placed on hospice

Weekends can heighten the risks around care transitions, as illustrated by the following three EMS responses:

> {a Friday response} Events prior to the incident: Pt refuses to eat or drink and has Hospice care scheduled for Monday morning but cant wait and wishes to be transported to {a hospital emergency department} for immediate hospice care.
>
> {a Monday response} [] has a history of [] and has been struggling with it for years and more so in last several weeks. [] was at another ER two days ago {a Saturday} for same problem, [] was in a hospice program for breathing
>
> {a Monday response} pt’s family states pt was released from [] hospital yesterday {a Saturday} after being admitted for [], family states hospice was not set up prior to pt being released from hospital

The following narrative, from an EMS response occuring on a Sunday, encapsulates all the issues of recent hospital discharge, early-stage hospice enrollment, and weekends. It also illustrates the distress felt by family members if they feel unsupported during a care transition:

> Dispatched to a [] {with difficulty breathing}. Upon arrival patient is responsive to verbal stimuli. Patient family states that patient was discharged from [] Hospital on Thursday of last week after a recent stay for []. According to family and paperwork provided patient was discharged home for comfort care and Hospice which was doing their intake on Monday of this week … The patient has not been taking [] medications, eating or drinking much for the last week. Throughout the night patient developed difficulty breathing, and restlessness. Family could not see the patient suffer like this and called 911.

#### 4.3.2 Emotions, culture, and confirmation of death

In 24 cases (14%), the patient was obviously dead at the scene. Of those 24 patients, 8 (33%) were early in their engagement with hospice care– “considering enrollment,” “intending to enroll,” “enrollment underway,” or “newly enrolled.” In most of these cases, the main function performed by the EMS crew, as documented in their narrative, was the confirmation of death, by means of physical examination and cardiac monitoring. Typical of these cases is the following:

> Pt was cyanotic and pulseless with lividity and rigor present. Applied cardiac monitor with initial rhythm of Asystole. Informed dispatch of obvious death and requested law enforcement. Upon law enforcement arrival, turned over custody of scene.

It was rarely clear from the narratives precisely why EMS was called in these cases. On occasion, difficulty contacting hospice or funeral home personnel for service in a timely manner was documented:

> Called to … [] Pt was on Hospice. Family was unable to get hospice on phone when Pt passed, so they called 911. Pt was checked on scene by medic []. Pt was not breathing, had no pulse, and was cold. Pt had valid DNR which is attached to PCR. Medical crew was released by deputies and returned to service.
>
> General impression was of … hospice pt with valid DNR, lying supine in bed, unresponsive and not breathing with family present. Pt last seen 2am. Family had contacted funeral home who in turn had her call 911.

Some interesting possibilities, purely speculative at this point, suggest themselves. They might open productive avenues of further research into richer emotional and cultural issues:

- Some survivors may doubt their ability to recognize death and derive reassurance from its confirmation by a health professional. By following societal expectations of calling 911 and hearing it from the first responders, it may also help loved ones to fully acknowledge and internalize the fact of the death.
- Scene narratives probably do not capture the full range of services provided by the EMS crew, and the first responders may have provided important sympathy and emotional support for the bereaved family.
- A number of recent hospice enrollees were among those obviously dead at the scene. It may be that these patients and families had not yet fully processed and embraced the fundamental shift in philosophy represented by hospice care, as described by some of Phongtankuel’s hospice staff focus group participants.^12^ Mentally and emotionally, they may still have been functioning in the customary medical, that is to say curative, model. From a young age, Americans are trained, by both formal and informal mechanisms, to call 911 in case of emergencies. This decades-long social learning may have overcome any anticipatory guidance provided to families in the process of hospice enrollment, resulting in reflexive calls to EMS. Examples of new hospice patients dead at the scene include:

> Dispatched to [] for a probable unattended. Upon arrival [], found [] also onscene. Directed to pt in hospital bed in bedroom of residence w/ first responder present. Pt was unconscious, unresponsive, pulsesless and apenic. First responder stated family had a DNR but could not find it. Pt’s family stated pt was placed on hospice care one week prior.

and

> Pt pulseless and apneic … Asystole in II, III, aVF … Spoke with [family member] who states pt just came home on hospice on Sat. States [] was tired all day and when [] checked on [] just prior to calling 911, [] wasn’t responding or breathing.

- All cultures have customs and rituals around death. These customs and rituals influence who is present at the time of death, what they do, and when and how they do it.^28,29^ One wonders whether ambulances, police cars, and uniformed first responders are part of a modern American death ritual that brings social and emotional support to the bereaved survivors during a “hyper-acute” postmortem period.

### 4.4 Advanced directives and negotiation of care

There was evidence of absent, unavailable, or ambiguous advanced directives or resuscitation wishes in 12 cases (7%). Ambiguity can be exacerbated in patients in early stages of the hospice enrollment process (see Section 4.3.1):

> Medical control contacted at [] ER and full pt care report given to Dr. []. Requested permission honor the family’s wishes given pt was to be a hospice pt and pt does not have a valid out-of-hospital DNR. Dr. [] agrees and orders resuscitation efforts to be withheld.

These ambiguities about resuscitation wishes sometimes led to likely futile, and perhaps undesired, medical interventions—something that hospice care is designed to prevent. The following example illustrates:

> Pt report from Paramedic [] states Pt is [] on Hospice Care at home with no DNR in place. Pt was noticed to be having severe respiratory distress this am by family members and 911 was called. Upon arrival of [] EMS, Pt in severe distress and alert to pain only. [] EMS on scene for several minutes due to family unable to determine if they wanted patient treated and transported. family finally agreed to have all that could be done for patient and transported to [] ED. Pt was removed from residence and went into respiratory arrest so Pt was intubated and bagged via BVM with additional help requested.

Even when written advanced directives are available, there can be issues of interpretation and application. In 25 cases (14.7%) there was documentary evidence of more than trivial or *pro forma* discussion, between two or more parties, of multiple management options that could be pursued. The parties most often involved were the EMS crew, the patient, family members, hospice personnel, the patient’s own physician, and the EMS medical control physician. The most common issues included 1. whether any EMS care at all was desired, 2. whether resuscitation would be attempted, and 3. whether the patient would be transported to the hospital.

These negotiations could sometimes be complex and time-consuming, as illustrated here:

> Hospice RN returns the call and goes over some options with the [family member] and pt. Ultimately the RN asks [family member] to contact the pt’s on call PCP {primary care physician} and ask them for advice. RN also advises to give the pt one of [] tablets. Pt is administered [] by [family member]. [] makes contact with pt’s on call MD, not[patient’s] normal PCP {primary care physician}. MD advises to send pt to the hospital without any real discussion of the pt’s problem. After the phone call the pt decides does not want to go to the hospital. EMS spends a long time talking with [family member] and assisting in the hospice process. The pt understands the situation and again refuses the hospital.

#### 4.4.1 Role of EMS providers

The above themes of emotion and culture, ambiguity of advanced directives, and negotiation of care raise important questions about the role and training of EMS providers. The original and still fundamental purpose of emergency medical services is to provide lifesaving care in urgent situations. But with an aging population with an increasing burden of chronic degenerative illnesses, resulting in frequent recurrent hospitalizations, yet often with shorter lengths of stay, additional training in these less technical, more psychosocial skills may be warranted. This would not be without challenges, chief among them the increased time and effort it would require of teachers and students, this in an era of increasing difficulty in recruiting and retention in the profession of EMS.,^30^ all of which has been exacerbated by the recent COVID-19 pandemic.

## 5 Discussion and conclusions

When patients are terminally ill, facing death within the next several months, they or their families often express a preference for a relatively non-interventional approach that emphasizes control of symptoms and that optimizes the patient’s sense of agency in managing their situation according to their wishes. Hospice programs are a valuable resource in this regard. Hospice patients, their family members, or their professional caregivers nevertheless sometimes call 911 (in the US) for EMS assistance. While the utilization of hospital emergency departments by hospice patients has been investigated, to our knowledge this is the first study to explore the contemporaneously-documented on-scene circumstances of *prehospital* care for patients enrolled or enrolling in a hospice program.

We found that EMS calls to the hospice patients in our sample arise generally through one or a combination of the following factors:

- Gaps in care during transitions, such as during the hospice enrollment process or after discharge from a hospital
- Perceived worsening of symptoms related to the patient’s “hospice-defining condition”
- Problems or symptoms unrelated to the patient’s “hospice-defining condition”
- Need for lift or re-positioning assistance
- Ambiguity about patient’s wishes, especially in regard to resuscitation
- Confirmation of death, perhaps ceremonial, ritualistic, or reflexive

Several of these finding are consistent with those reported by interview-based studies. Chief complaints are similar to those itemized by Phongtankuel et al,^13^ with respiratory distress, altered mental status, pain, and falls being prominent. The risk to patients inherent in transitions from one type or location of care to another is again confirmed here. Others, including the frequency of lift-assist and confirmation of death, have not previously been described. Summoning EMS to confirm that a hospice patient has died is a particularly interesting phenomenon that warrants further study, in that it seems an inefficient use of already overburdened EMS resources.

EMS responses to hospice patients by no means represent necessarily a poor outcome or are uniformly undesirable. Some are precipitated by situations that would reasonably warrant emergency medical care in anyone, enrolled in hospice or not. Others (for example, lift assistance) are valuable in that they help restore a patient or a family to a stable biopsychosocial state with minimal intervention and without disturbing the trajectory of their hospice care with an ED visit. However, some lead to a cascade of medical interventions,^11^ including possibly transport to a hospital, that the patient and family may, given other resources or options, have wished to avoid. A better understanding of that interface might illuminate changes that could be made to improve it, with a goal of ensuring that hospice patients and their families receive emergency medical services when they need and can benefit from them but receive other, non-EMS, services when those are more suitable to their needs.

### 5.1 Limitations

This was a retrospective study, conducted via review of electronic patient care reports that were generated as part of standard EMS operations. There was no purpose-designed, prospective data collection instrument. Emphasis was on exploring themes found in EMS provider narratives. The free-text nature of the provider narrative in NEMSIS make it potentially a rich source of detailed information. Conversely, that free-text nature can make the narratives highly variable in their information content. The absence from the narrative of documentation of a feature or action at the scene does not necessarily mean it was not present or undertaken.

It is possible that any given patient is represented more than once in the analytical dataset, either because two different EMS agencies responded together to the call (“tiered response” with both BLS and ALS) or because EMS was called to assist the same patient on more than one occassion during the study period.

The experiences of one EMS system and two hospice agencies in one small geographic region may not be generalizable. On the other hand, this is the only in-depth, contemporaneous study of EMS responses to patients in hospice of which we are aware, and we believe the insights gained are valuable and can stimulate similar research in other settings.

We did not observe the care at the scenes, nor did we interview patients, family members, or EMS providers. Thus it is difficult to draw conclusions about the thoughts, feelings, or motives underlying any of their actions. That ground, however, appears to be well-covered in the existing literature.

## Data Availability

Due to privacy concerns, the orginal data cannot be made available.

